# Rare variants and founder effect in an understudied Quebec population

**DOI:** 10.1101/2025.01.21.25320907

**Authors:** Mylène Gagnon, Claudia Moreau, Jasmin Ricard, Marie-Claude Boisvert, Alexandre Bureau, Michel Maziade, Simon L. Girard

## Abstract

Founder events influenced the genetic diversity within the Quebec province, increasing the frequency of certain rare pathogenic variants in regional populations. Some regions such as Beauce remain understudied despite evidence of a regional founder effect. Leveraging extensive genealogical data, we found a specific regional structure emerging in Beauce following the initial settlement, with a gradual increase of the inbreeding and kinship coefficients and a low ancestors’ diversity. Profiting from the genetic distinctiveness of the region, we identified 28 rare pathogenic variants with higher carrier rates in Beauce than in urban regions due to its regional founder effect. This provides the first in-depth study of Beauce’s genetic and genealogical landscape, revealing a distinct structure and suggesting that other overlooked regions, in Quebec and elsewhere, could benefit from fine-scale population structure study to improve the understanding and management of rare diseases.

## 1 Introduction

The genetic specificity of populations with founder effects provides a valuable perspective to identify rare variants as their genetic isolation can lead to an increase or a decrease in the prevalence of rare pathogenic variants^1–3^. A good example is the province of Quebec, which has experienced both founder effects and genetic drift. These phenomena contributed to an increased frequency of certain alleles within the population, but regional variations in the prevalence of those alleles can also be observed^4^. Approximately 74% of the almost 9 million present-day Quebecers are of French-Canadians origin, descending from the 8,000-10,000 French settlers who embarked on the settlement of New France between 1608 and the British Conquest of 1759^5–7^. While the province did not adopt a policy of outright exclusion towards immigration, the incoming subsequent waves of immigrants of diverse origins had a limited genetic impact on the established population^8^. The principal driver behind the population expansion was the pronounced natality rate. The accelerated population growth initiated a cascade of effects, including the territorial expansion of Quebec and the opening of settlements in regions previously unoccupied by European settlers^9^. The establishment of settlements in remote and geographically isolated areas further contributed to the subdivision of the population and engendered a spatially fragmented genetic structure, with distinct regional populations within the province^10,11^. This laid the foundation for the distinctive genetic makeup characterizing present-day Quebecers of French descent.

An illustration of a distinct subpopulation within Quebec considerably diverging from the major urban centers is the region of Saguenay–Lac-St-Jean (SLSJ). While SLSJ’s genetics have been extensively studied^12–15^, some regions, presumed to be minimally affected by demographic processes, have been overlooked. One such region is Beauce, situated in Chaudière-Appalaches (Figure 1). Beauce’s settlement began in 1736 with Europeans primarily arriving from areas near Quebec City. Immigration mostly occurred in families, as for SLSJ, and the fertility in Beauce was high, with rates similar to those observed across the province, with the exception of SLSJ^16^. The Beauce region stayed open to immigration, but was difficult to access, contributing to its isolation. Settlement first started not far from Quebec City and gradually extended to further territories^17^. While this territorial expansion was primarily driven by local settlers, European immigrants also contributed. However, the proportion of the population with non-French-Canadian descent remained low and only decreased over time, dropping to less than 2% by 1931^16^. Despite those characteristics indicating a regional founder effect, the genetics of Beauce remain understudied. Current knowledge of the region’s genetic landscape is limited to findings derived from simulated genomic data or clinical observations of rare disease patients^18–20^. Notably, autosomal recessive cerebellar ataxia type 1 (ARCA1), also known as ataxia Beauce type, was first described in the region in 2007 and is mostly seen in the French-Canadian population^20,21^.

**Figure 1.**
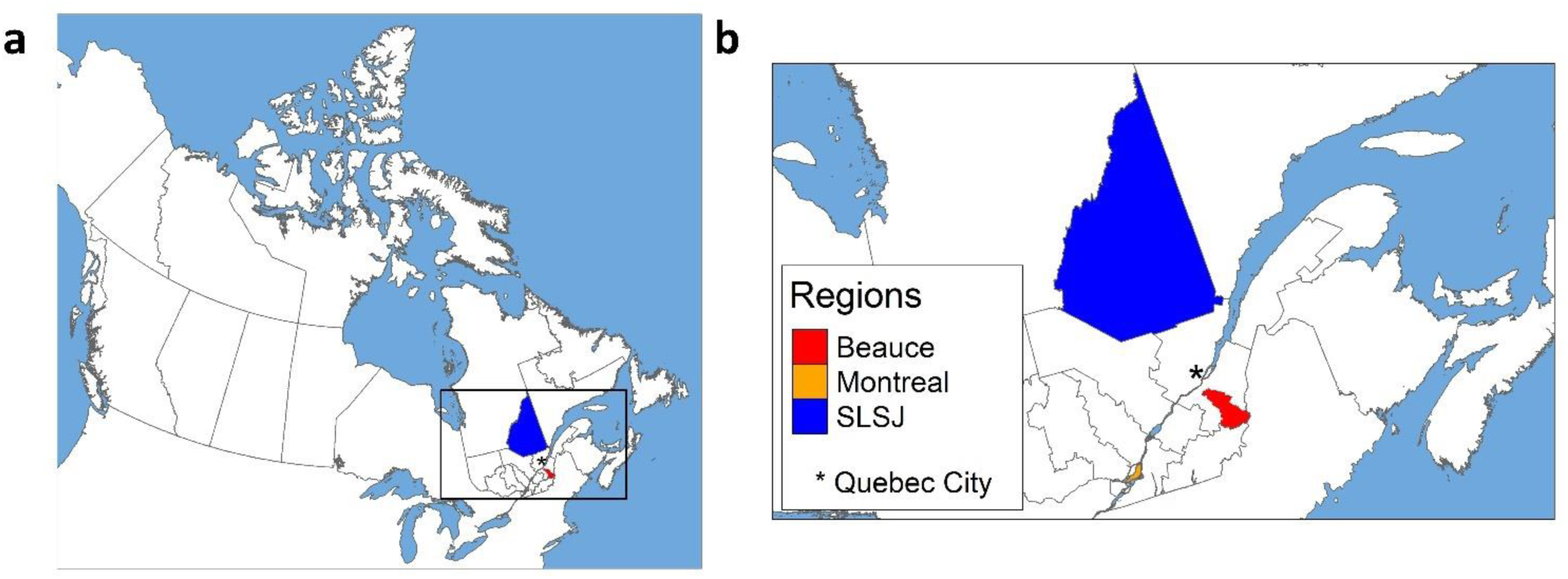
Map of **a** Canada and **b** southern Quebec highlighting the areas of relevance to this study.

A global portrait of the prevalence of rare variants in Beauce has never been traced, likely due to the underrepresentation of rural areas in Quebec’s genetic cohorts. Therefore, this study aimed to characterise the genetics of Beauce’s population and to investigate how its structure can be explained by the demographic processes using deep genealogies. We observed that Beauce exhibited heightened kinship and inbreeding coefficients and a low ancestors’ diversity. Then, leveraging the genetic distinctiveness of the region, we looked at the frequency of rare pathogenic variants and identified 28 variants that are more frequent in Beauce due to a regional founder event.

## 2 Subjects and methods

This study was approved by the *Université du Québec à Chicoutimi* and the *Centre Intégré Universitaire de Santé et de Service Sociaux de la Capitale-Nationale* ethics boards. Written informed consent was obtained from all adult participants.

### 2.1 Genealogical data and cleaning

The genealogical data used in this study was obtained from the BALSAC database^22^. BALSAC is a repository containing Catholic marriage records and civil registry data used to reconstruct genealogies. The available data ranges from the beginning of the European settlement in the early 1600s to 1965.

To describe the regional structure of the populations, we selected all individuals married in Beauce, SLSJ and Montreal between 1935 and 1960 and reconstructed their genealogies going back over three centuries, thus covering almost the entirety of Quebec’s post-European settlement history. We then performed a quality control based on the completeness of the genealogies which corresponds to the proportion of known ancestors on all expected ancestors for every generation^23^. For Beauce and SLSJ, only genealogies with a perfect completeness up to the third generation were included. For Montreal, since the overall completeness considerably dropped after the fourth generation, only the genealogies complete to the fifth generation were retained. All three regions then showed similar completeness throughout generations (Figure S1). Then, we removed first-degree relationships among the probands to avoid redundancies and, to ensure that the genealogies accurately reflect the regional ancestry, probands who immigrated recently (within three generations) were removed. The final dataset is described in Table 1.

**Table 1.** Description of the genealogical data for each regional group.

| Region | Number of probands | Number of ancestors |
| --- | --- | --- |
| Beauce | 7,445 | 61,569 |
| Montreal | 10,048 | 239,011 |
| SLSJ | 9,655 | 78,714 |

### 2.2 Genealogical analyses

All genealogical analyses were conducted with the Julia package GenLib.jl version 0.1.4^24^. For each regional group, we measured the evolution of the averaged kinship and inbreeding coefficients, as well as the ancestors’ diversity ratio (ADR) for each decade from 1640 to 1950. Kinship being directly influenced by the presence of close relatedness between pairs, relationships of the first degree were removed for each decade.

We also assessed the ADR to represent the concentration of the ancestors in each regional group. To measure the ADR, we computed two metrics: the number of ancestors present in the genealogy of all probands, and their occurrence. The occurrence represents the overall count of all ancestors identified, including multiple apparitions, while the number of ancestors reflect the distinct individuals counted without repetition^25^. We then computed the ratio of the number of ancestors to the occurrence of ancestors as follow:

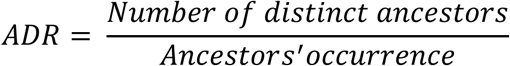

To ensure comparable measurements between the regional groups, we used a bootstrap approach where we randomly selected a subset of 7,445 probands from each regional group. The replicated sample size corresponds to the number of probands in the smallest regional group. This process was repeated 1,000 times, and we estimated the minimum, the maximum, and the mean ADR.

### 2.3 Genotyping data and cleaning

The genetic data in this study includes the CARTaGENE cohort (CaG), and the Eastern Quebec Schizophrenia and Bipolar Disorder Kindreds cohort (SZ-BP). The CaG cohort comprises randomly selected individuals residing in metropolitan areas in the Quebec province^26^. The SZ-BP cohort comprises 48 multigenerational families affected by schizophrenia and bipolar disorder. It includes 1,200 individuals recruited in Beauce, SLSJ, and Iles-de-la-Madeleine^27,28^. The content of both samples is described in Table 2.

**Table 2.** Description of the genomic data available.

| Cohort | Recruitment region | Data type | Sample size |
| --- | --- | --- | --- |
| CARTaGENE | Gatineau, Montreal, Quebec City, SLSJ, Sherbrooke and Trois-Rivieres | Genotyping | 28,337 |
|  |  | Whole genome sequencing | 2,184 |
| Eastern Quebec Schizophrenia and Bipolar Disorder Kindreds | Beauce, SLSJ and Iles-de-la-Madeleine | Genotyping | 1,200 |
|  |  | Whole Genome Sequencing | 500 |

Full description of the genotyped data from the CaG cohort is available in its documentation (https://cartagene.qc.ca/files/documents/other/Info_GeneticData3juillet2023.pdf). For the SZ-BP cohort, genotypes were carried out in two waves using DNA extracted from immortalized lymphocytes or fresh blood by affinity column (Midi prep Qiagen). 585 subjects were genotyped at 622,184 autosomal SNPs with the Illumina Infinium Human OmniExpress array and 615 subjects were genotyped at 502,425 SNPs with the Illumina Global Screening Array, more details are provided in Bahda *et al*^29^.

Each genotype dataset was cleaned independently, then merged using plink/1.9b_5.2-x86_64^30^. SNPs missing in at least 5% of the subjects were excluded, and individuals with a genotype missingness of at least 5% were removed. SNPs that do not conform to the Hardy-Weinberg Equilibrium with a P-value threshold of 10^-6^ were also removed. Cleaned datasets were then merged based on the intersection of positions. After the merge, the same quality control criteria were reapplied, and alleles with a minor allele frequency (MAF) below 5% were excluded. The final dataset comprises 29,448 individuals with 132,700 SNPs.

### 2.4 Ancestry-based clustering

Since individuals in the CaG cohort were recruited according to their residing region instead of their birthplace, we formed population clusters based on their genetic ancestry. To do so, a principal component analysis (PCA) was performed on the genotype data using plink. Because of the high proportion of relatedness in the SZ-BP cohort, we chose not to exclude related individuals. A classical PCA method was chosen instead of a projection of the related over the unrelated individuals to avoid the shrinkage bias associated with this method, which is especially pronounced when the unrelated sample size is limited^31^. Furthermore, the shrinkage is increased when computing principal components (PC) that explain lower variance and those were needed to assess the fine structure of the population, as Beauce became distinct from the other Quebec regions on the 10^th^ PC. Using the first 10 PCs, a uniform manifold approximation and projection (UMAP) was performed with the R umap/0.2.10.0 library^32^. The 10 first PCs were chosen based on the third cutoff point identified in the scree test and because it allowed a visual separation of individuals recruited in Beauce (Figure S2-S3). The UMAP was performed to emphasize the local genetic structure within the data and was done on the 10 first PCs instead of the genotyped data to remain computationally efficient^33^.

Density-based spatial clustering of applications with noise (DBSCAN) was performed on the UMAP projection using the dbscan library^34^ to identify our ancestry-based clusters^35,36^. The size of the epsilon neighborhood was set to 0.30, and the minimum number of points in the neighborhood was set to 30. Those parameters were chosen to ensure the identification of meaningful clusters while minimizing the inclusion of noise.

This resulted in 18 distinct clusters, and their origin was assigned based on the recruitment region or the birthplace of most individuals within the clusters. For individuals born outside of Canada, we used the auto-reported country of birth of the individuals and grouped them in broader categories. Clusters one and four were kept for further analysis. They respectively corresponded to Beauce, and the urban regions of Quebec (UrbanQc), and will be referred to as such for the subsequent analyses. This left us with 16,553 genotyped individuals. The Beauce cluster comprised 752 individuals of which 65% were actually recruited in Beauce. The rest were recruited mostly in metropolitan areas without information about their birthplace. The UrbanQc cluster comprised 15,801 individuals recruited mostly in urban centers (Figure S4).

### 2.5 Identification of rare pathogenic variants more frequent in Beauce

To identify rare variants with higher frequencies in the Beauce cluster, we used the whole genome sequencing (WGS) data available for some individuals in the SZ-BP and CaG cohorts (Table 1). For the SZ-BP cohort (n = 500), we used the DRAGEN Germline Pipeline v3.10.4 on the Illumina BaseSpace to align the FASTQ on the hg38 Human reference genome and to call variants^37^. For the WGS of CaG (n = 2,184), details are available in the documentation (https://cartagene.qc.ca/files/documents/other/Info_GeneticData3juillet2023.pdf). The gVCF files from the SZ-BP and CaG cohorts were joint called using the Population explorer WebApp (https://popex.dragen.illumina.com/) to produce a joint VCF file. WGS are available for 223 individuals in the Beauce cluster, and 1,092 individuals in the UrbanQc cluster.

We selected variants in the ClinVar database^38^ (version of June 24th 2024) following the same criteria as Michel *et al.*^39^, leaving a total of 240,716 variants. We determined the MAF of the selected variants in the Beauce and UrbanQc clusters using plink. Since most of the individuals in the Beauce cluster are from the SZ-BP cohort (n_CaG_ = 26, n_SZ-BP_ = 197), we compared the MAF of all the rare pathogenic variants between the cohorts to ensure the absence of a batch effect (Figure S5). To identify the variants at higher frequencies in Beauce, we calculated the relative frequency difference, which corresponds to the relative difference between the MAF. The relative frequency difference is calculated as follow:

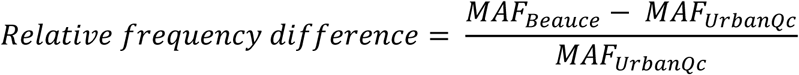

Only variants present in at least five carriers and reaching a relative frequency difference of at least 10% in the Beauce cluster compared to the UrbanQc cluster were considered. A liberal threshold of 10% was chosen as suggested by Michel *et al.*^39^, allowing to minimize the error of results omission. MAF were also obtained for non-Finnish Europeans from gnomAD v4.1.0^40^. Because our study focuses on rare variants, three variants with a MAF exceeding 5% were excluded. Finally, carrier rates were calculated using the proportion of heterozygous individuals for each variant in the Beauce and UrbanQc clusters. Heterozygous individuals were identified using plink.

### 2.6 Classification of rare pathogenic variants more frequent in Beauce

We used identical-by-descent (IBD) segments shared between the carriers of each variant to identify the variants that are more frequent in the Beauce cluster due to a regional founder effect. IBD segments were inferred on the phased genotypes with refinedIBD version 17Jan20^41^ within Beagle version 18May20^42^. The IBD segments were detected using a sliding window of 40 SNPs, a log of odds threshold of 3, and a minimal length of 2 cM with a sensitivity scale of 10 to retain segments in familial data. Segments were then merged using the merge-ibd-segments 17Jan20.102 tool^41^. For each enriched variant, we examined the IBD sharing between the carriers and determined the proportion of pairs sharing at each genomic position. We considered the variant as founder if at least 50% of the carriers’ pairs were sharing IBD around the variant’s position. To ensure that the variants were indeed linked to a founder effect and not biased by the related individuals present within the sample, we inferred relationships between the carriers using the total of IBD sharing. Variants of familial origin were defined as variants carried by less than five individuals related further than the third degree (proportion of sharing ≤ 0.125). Variants that did not fit either category were classified as variants with multiple introductions in the population. The steps for the identification of the founder variants are summarized in Figure 2.

**Figure 2.**
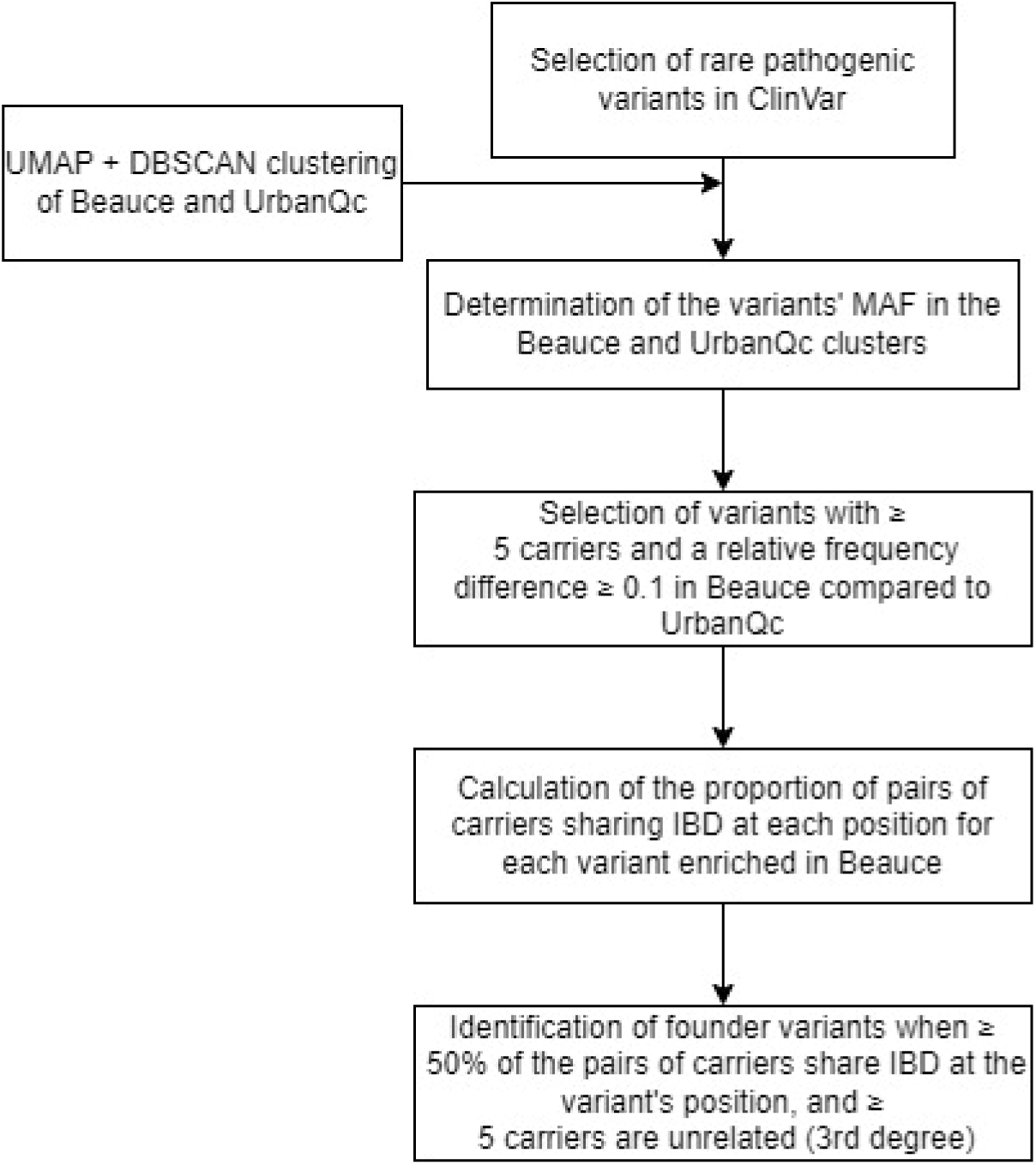
Overview of the founder variants’ identification strategy.

## 3 Results

### 3.1 Genealogical regional structure

The evolution of the regional population genealogical structure of Beauce, SLSJ, and Montreal were compared using the mean kinship, and inbreeding coefficients (Figure 3). In Beauce and SLSJ, before 1820, the kinship increased gradually while the increase in inbreeding was steeper, reflecting the early stages of settlement where a limited number of founders had a significant contribution exacerbated by the regional isolation. However, Montreal displayed considerably lower values during this period, likely due to its larger and more diverse set of founders. After the settling of SLSJ, both coefficients kept increasing drastically until they started to stabilize. This plateau had been observed in previous studies of the SLSJ population structure^10^. This stabilization is similarly observed in Montreal, while in Beauce, the inbreeding coefficient continued to rise steadily.

**Figure 3.**
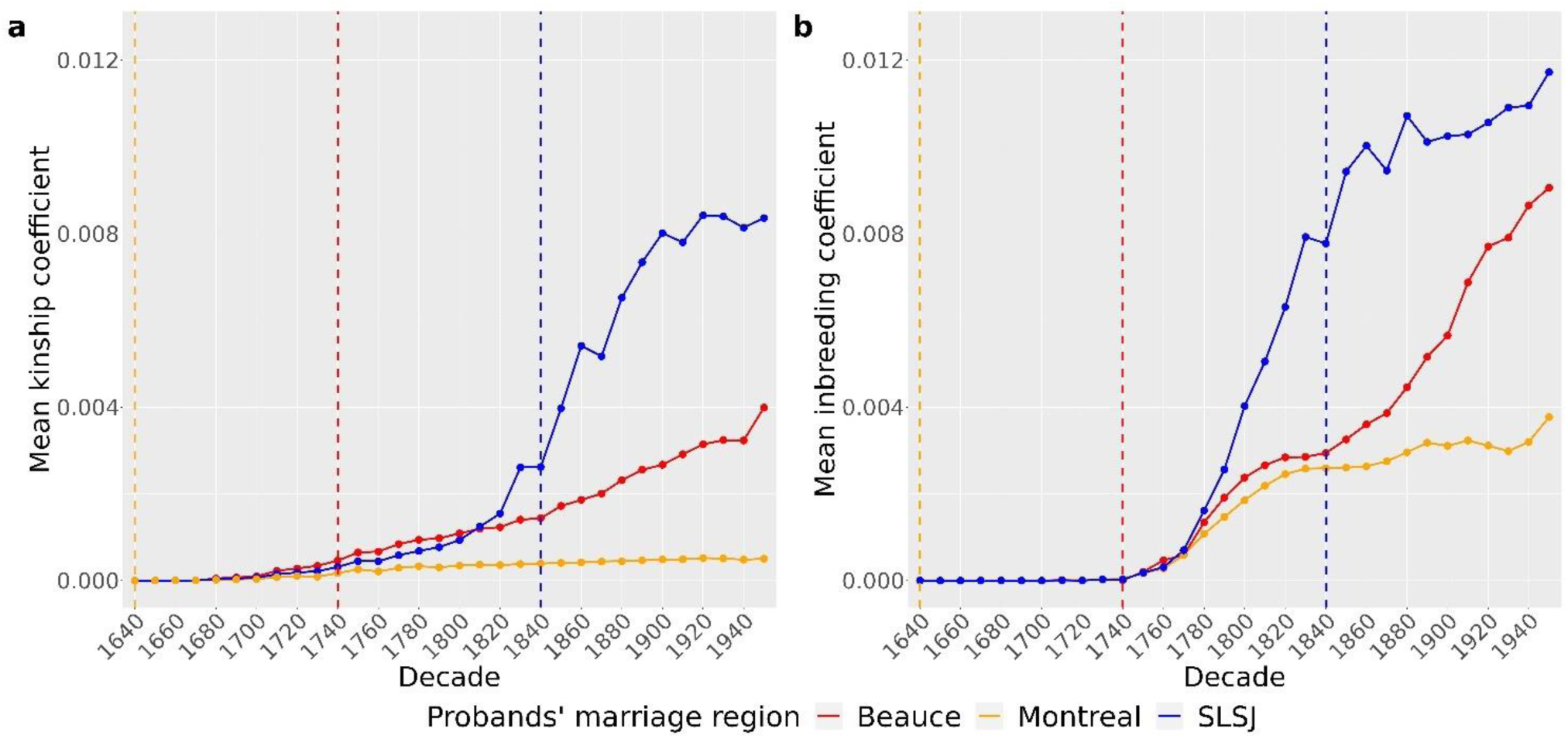
Average **a** kinship and **b** inbreeding coefficients of ancestors of each regional group per decade. Vertical dotted lines represent the decade of the beginning of the settlement for each region.

Across the three regional groups, the inbreeding coefficient consistently exceeds the kinship coefficient, as the kinship coefficient is calculated between each pair of individuals whereas the inbreeding coefficient represents the kinship between the pairs that effectively reproduced. Interestingly, the difference between the kinship and inbreeding coefficient is the lowest in SLSJ. In 1950, SLSJ inbreeding is 1.4 times higher than its kinship, while in Beauce, the inbreeding is almost 2.3 times the kinship.

We then measured the ADR at each decade for the three regional groups as an indicator of the ancestors’ concentration (Figure 4). The number of distinct ancestors gradually increased over time for each region. Montreal consistently shows the highest diversity, reflecting its position as a major urban center. In contrast, Beauce and SLSJ show considerably lower ADR throughout most of the observed period. The low ADR in Beauce is driven by the relatively frequent occurrence of a broad set of ancestors, whereas in SLSJ, it is driven by the presence of a few ‘super-ancestors’ who appear at an exceptionally high number of times (Figure S6).

**Figure 4.**
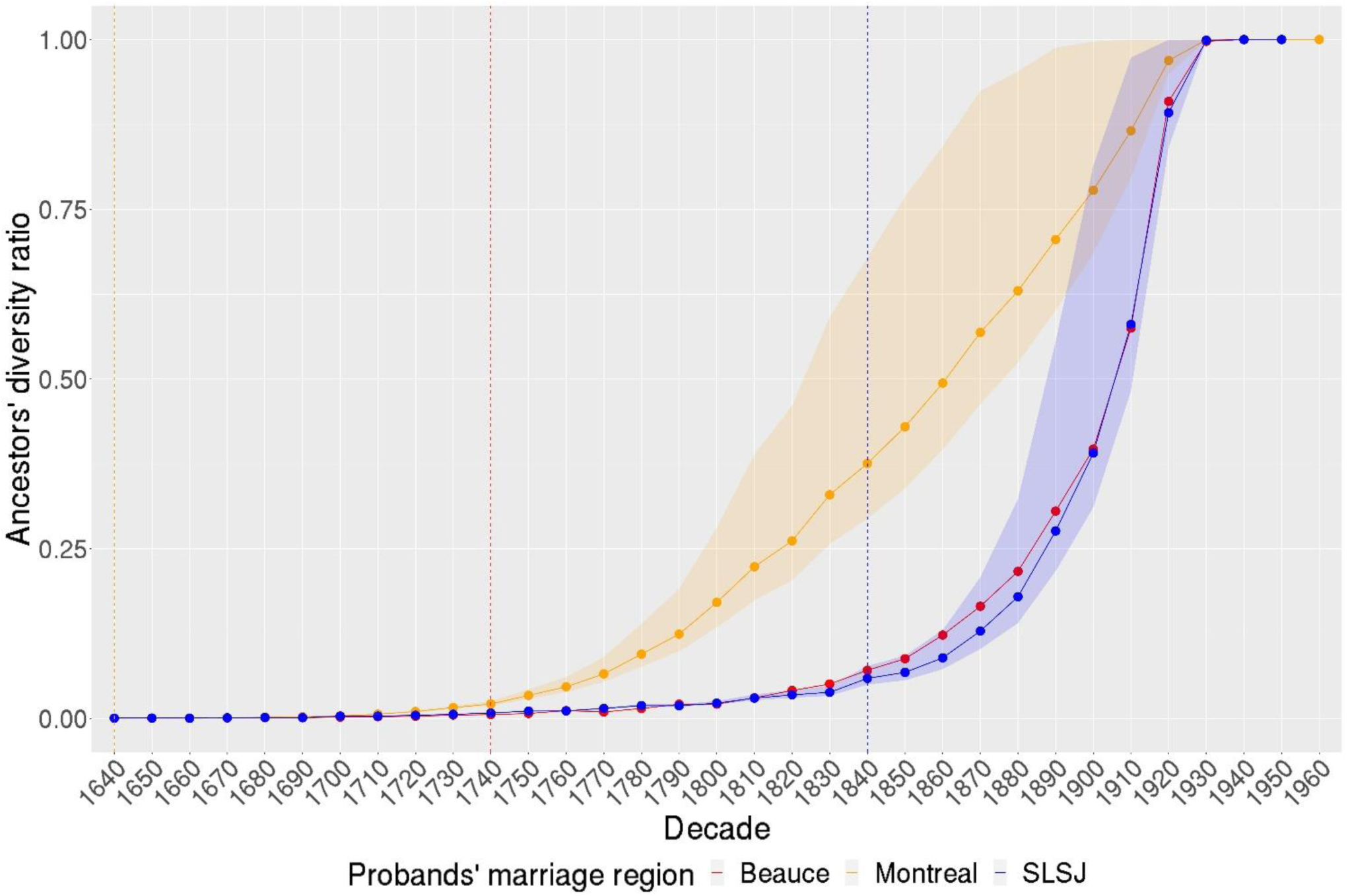
Ancestors’ diversity ratio of each regional group per decade. Ancestors were bootstrapped 1,000 times for 7,445 probands and intervals correspond to the minimum and maximum values while the full lines represent the mean. Vertical dotted lines represent the decade of the start of the settlement for each region

### 3.2 Regional genetic structure

The present-day population structure of Quebec and its regions was assessed using PCA (Figure S3) and UMAP (Figure 5) on the genotype data to identify clusters based on ancestry. These analyses revealed distinct regional clusters, including clusters associated with recent immigrants born outside of Canada. Cluster one, mostly composed of individuals from the SZ-BP cohort recruited in Beauce, was entirely separated from the fourth cluster, the main one representing metropolitan Quebec areas. Cluster five, associated with SLSJ ancestry was also distinguishable from the other regional populations, as expected from previous studies^1,18,39^.

**Figure 5.**
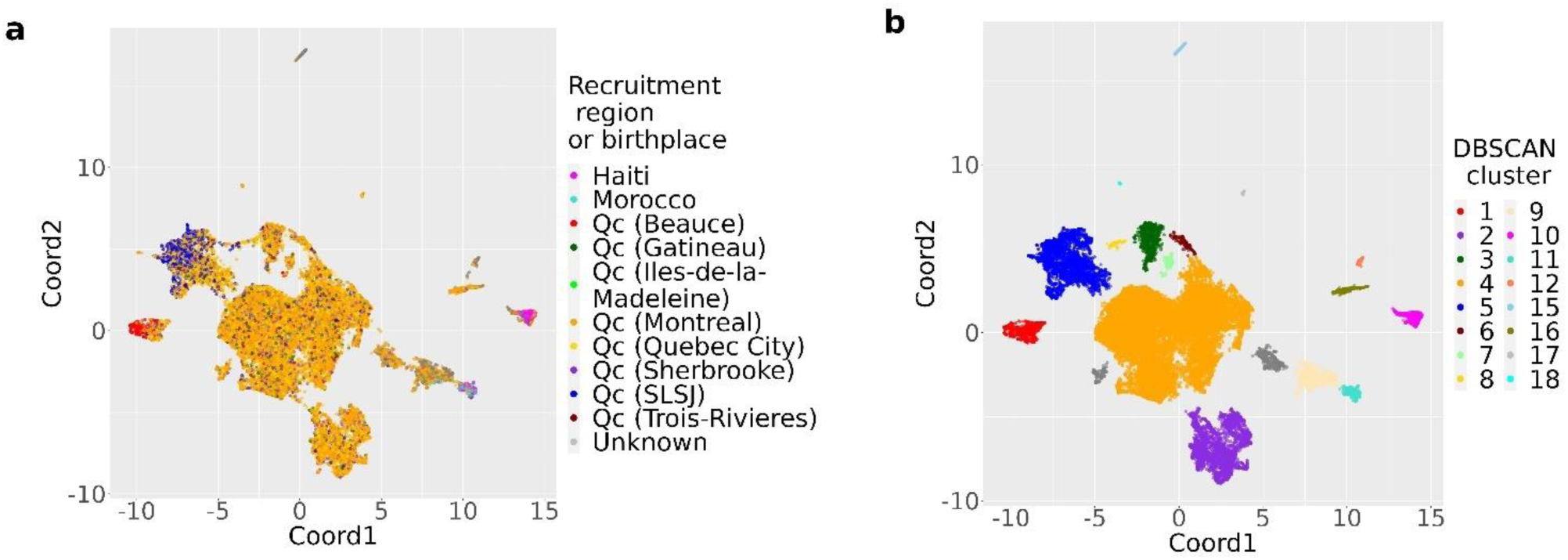
UMAP of the first 10 principal components of the genotype data colored by **a** recruitment region or birthplace if outside of Canada and **b** DBSCAN cluster.

DBSCAN identified 18 distinct clusters while excluding outliers, thereby refining the population structure. We inferred the ascendance based on the recruitment region or birthplace of the individuals within the clusters. Multiple clusters were associated with metropolitan areas; however, only the largest of these clusters was retained for analysis. Those smaller clusters may represent more admixed individuals or undefined regional populations.

### 3.3 Rare pathogenic founder variants

Among the WGS of 223 individuals in the Beauce cluster, we identified 32 rare pathogenic variants with a relative frequency difference of at least 10% compared to the UrbanQc cluster. For most variants, carriers were found in both cohorts (CaG and SZ-BP) within the Beauce cluster (Figure S7). For 28 variants, more than half of the carriers were sharing IBD around the variant’s position and were therefore described as founder variants in the Beauce cluster (Table 3 and Supplementary Data 1). The remaining four likely arose from multiple introductions as assessed by the insufficient IBD sharing among the carriers (Table S1 and Figure S8).

**Table 3.** Rare pathogenic founder variants with a relative frequency difference of at least 10% in Beauce compared to UrbanQc and a carrier rate higher than 1/25.

| HGVS name | Disease (ClinVar ID) | Inheritance | MAF NFE | CR Beauce | CR UrbanQc |
| --- | --- | --- | --- | --- | --- |
| NM_000022.4:c.956_960del | Severe combined immunodeficiency (193544) | AR | 0.0002 | 1/14 | 1/273 |
| NM_001199251.3:c.67A>G | Chronic atrial and intestinal dysrhythmia (162627) | AR | 0.0002 | 1/15 | 1/99 |
| NM_015665.6:c.856C>T | AAA syndrome (1322889) | AR | 0.0001 | 1/16 | 1/273 |
| NM_001698.3:c.656-2_656-1del | 3-methylglutaconic aciduria type 1 (1067674) | AR | 0 | 1/16 | 1/546 |
| NM_020458.4:c.1001+3_1001+6del | Gastrointestinal defects and immunodeficiency syndrome type 1 (50608) | AR | 0 | 1/19 | 1/364 |
| NM_138694.4:c.6793C>T | Autosomal recessive polycystic kidney disease (1946278) | AR | 0 | 1/22 | 1/156 |
| NM_182961.4:c.15918-12A>G | Autosomal recessive cerebellar ataxia type 1 (2326) | AR | 0 | 1/22 | 1/273 |

AD: Autosomal dominant, AR: Autosomal recessive, CR: Carrier rate, MAF: Minor allele frequency, NFE: Non-Finnish Europeans

Carrier rates in Beauce varied from 1/14 to 1/45 and the highest carrier rate observed in UrbanQc is 1/57 for the variant NM_001048174.2:c.1103G>A, which also exhibits the highest MAF among non-Finnish Europeans from gnomAD. Two variants were virtually absent from UrbanQc while exhibiting carrier rates of 1/32 and 1/45 in Beauce.

## 4 Discussion

### 4.1 Regional population structure characterisation

The first objective of this study was to characterize the population structure of Beauce using extensive genealogical data. Our findings provide insight into how the demographic history of Beauce has shaped its genetic profile, leading to a clear differentiation from the urban areas of Quebec. This structure is evident in the kinship and inbreeding coefficients in Beauce, which gradually increased following the initial settlement in 1736. For Beauce and SLSJ, settlers primarily originated from a small number of founding families coming from surrounding areas, leading to heightened kinship, but their demographic history diverged. We observe a pre-settlement structure emerging in SLSJ before 1840, while most of the ancestors were located in Charlevoix^10,43^. SLSJ then experienced a rapid demographic expansion resulting in elevated kinship and inbreeding coefficients^10,11^. In contrast, Beauce’s population growth was more gradual with a fertility rate comparable to what was observed in the other regions of Quebec^16^. The relative geographic isolation of Beauce still led to elevated kinship and inbreeding coefficients since marriage patterns were largely confined to the community due to limited immigration^17,44^. In regions where gene flow is limited, there is a direct relationship between time and distant inbreeding^45^. Over time, such isolation can result in a reduced genetic diversity within the population^46^, as seen in Beauce, where an increasing number of common ancestors among individuals led to an increase in kinship. As individuals with increasingly closer kinship remain in the region and reproduce together, the inbreeding coefficient rises gradually^47^. Unlike SLSJ and Montreal, where inbreeding stabilized over time, Beauce saw a continued increase throughout the observed period. With a population less than half the size of SLSJ^48^, individuals in Beauce were more likely to reproduce with distant relatives, contributing to the ongoing rise in inbreeding coefficients.

While inbreeding is generally higher than kinship in every regional population of Quebec, the difference between those measures is greater when closely related individuals produce a descendance^49^. The higher the kinship is between the parents of an individual, the higher their inbreeding coefficient will be^23^. After the end of the 19^th^ century, there is a larger disparity between the kinship and inbreeding coefficients in Beauce and Montreal compared to SLSJ. Beauce saw a higher rate of reproduction among closer ancestors compared to SLSJ. Vézina *et al.*^25^ already established that the close inbreeding in SLSJ is the lowest in the province, while the eastern regions like Beauce show the highest close inbreeding coefficients. In Montreal, even if the overall kinship and inbreeding are the lowest in the province, the close inbreeding is almost twice as high as in SLSJ^25^, which explains why there is a greater disparity between the inbreeding and kinship coefficients than in SLSJ.

Despite differences in their demographic processes, Beauce and SLSJ share similar trends, such as low ADR throughout the observed period of the present study. The low ADR observed in those regions indicates origin from a limited number of initial settlers and limited interregional admixture, increasing the likelihood of shared ancestry among individuals. This can partly explain the high kinship coefficient of Beauce and SLSJ observed in Figure 3. In contrast, we found a high diversity of ancestors in Montreal, where constant immigration brought individuals not only from other regions of Quebec but also from other countries^50,51^. The ancestors’ diversity of the regions has been inferred in the previous studies through the distribution and frequency of family names as both are known to correlate^46,49^. Bouchard *et al.*^52^ analyzed the relative frequency of the most common family names in each region to estimate ancestral diversity. Their findings revealed that Montreal exhibited the highest ancestral diversity, while SLSJ and Beauce ranked lower, indicating reduced diversity. This was also reflected by the depth of regional ancestral roots which represents the number of generations needed to find a majority of ancestors coming from outside of the region^44^. The low ADR we observed in Beauce is consistent with its high intraregional kinship and the gradual increase in its inbreeding coefficient, highlighting a population structure with significant shared ancestry.

The measure of ADR is largely influenced by the occurrence of the ancestors, which is driven by the demographic expansion process. The population growth in SLSJ was five times greater than in the other regions of Quebec, including Beauce^16,53^. Being on the front of this rapid wave of expansion conferred a high reproductive success to the initial settlers of SLSJ, who left an important number of descendants^54^. The reduced ADR in SLSJ reflects this demographic pattern. Few ancestors have extremely high occurrences, while the rest appear at low frequencies (Figure S3). In contrast, the gradual localized population growth in Beauce led to a different pattern, where most ancestors exhibit consistently high frequencies, resulting in a similarly reduced ADR. This also resulted in an elevated kinship in Beauce, as individuals share a high number of common ancestors. While elevated, the kinship and inbreeding coefficients in Beauce are still lower than what is seen in SLSJ because of the more gradual population expansion.

As previously demonstrated, the genealogical structure of a population mirrors its genetic structure^10,11,18^. Thus, the high kinship and inbreeding coefficients and the low ADR in Beauce are indicative of a population where individuals share high proportions of their genomes. Rare pathogenic variants could have been introduced by initial founders, then spread through drift in the population, leading to a higher frequency than what can be observed in populations that did not go through this regional founder effect.

### 4.2 Rare pathogenic founder variants in Beauce

The second aim of this study was to explore the frequency of rare pathogenic variants in Beauce compared to more admixed regions to identify variants present at higher frequencies. More precisely, we wanted to identify variants with a higher frequency in Beauce than in the metropolitan areas of Quebec because of the regional founder effect.

We first defined population groups using a UMAP visualization to identify compact clusters of individuals with similar genetic backgrounds^33,35^. The distinct regional clusters demonstrate that the Quebec population cannot be treated as a single homogeneous entity, as it comprises multiple, regionally defined subpopulations with unique genetic and demographic histories. This clustering enhanced the identification of rare pathogenic variants concentrated in specific regional groups, where their increased frequency facilitated detection^1,39,55^. Using this method, we identified 32 rare pathogenic variants with a relative frequency difference of at least 10% between the Beauce and UrbanQc clusters. In addition to accounting for their high frequency, we examined the IBD segments sharing between carriers and identified 28 rare founder pathogenic variants in Beauce.

Among those, 14 were previously reported in Quebec. Notably, NM_182961.4:c.15918-12A>G was identified as founder in Beauce, exhibiting a carrier rate of 1/22. In Eastern Quebec, it is the most frequent of the seven ARCA1 variants reported among individuals of French-Canadian descent, with a minimum carrier frequency reported at 1/134^20,56^. Since previous studies focussed on a broader region, they obtained a much lower carrier rate than what we observed. Including metropolitan areas in their study likely diluted the frequency of the variant because, as shown in the present study, this variant is around 12 times more frequent in Beauce than in urban populations of Quebec. This elevated frequency was previously attributed to a founder effect in the French-Canadian population based on the prevalence of the disease and genealogical reconstruction^20^ and our analysis of IBD sharing confirmed its founder origin in Beauce. No other known variants for ARCA1 were found in the Beauce cluster, but three were present at low MAF in UrbanQc.

For two of our previously reported founder variants, the IBD sharing between the carriers in Beauce and UrbanQc indicates a shared founder event. NM_001199251.3:c.67A>G had already been suspected to be more frequent among French-Canadians due to a founder effect^57^ and NM_138694.4:c.6793C>T was reported as founder in SLSJ with a carrier rate of 1/163^39^. We observe it at a much higher frequency in Beauce, with a carrier rate of 1/22. The other previously described founder variants were only founder in Beauce. For NM_020458.4:c.1001+3_1001+6del, a variant associated with gastrointestinal defects and immunodeficiency syndrome type 1, a founder effect was suspected among French-Canadians because of its high frequency^58^. It was also described as founder in SLSJ, with a carrier rate of 1/56^39^. Again, we observed a higher carrier rate with 1/19. We also described a variant associated with Tay-Sachs disease, NM_000520.6:c.1274_1277dup, to be more frequent in Beauce because of the founder effect. Multiple Tay-Sachs variants are reported in Quebec, with regional differences in frequency. While our founder variant was previously described as more frequent among French-Canadians, it was never traced back to a specific region^59^.

A recent study in SLSJ identified 80 rare founder pathogenic variants and approximately half were previously documented^39^. Unlike SLSJ, Beauce does not benefit from extensive genetic data and a well-documented prevalence of rare diseases. By applying a similar methodological approach to Beauce, we provide reliable findings despite the limited literature on the prevalence of genetic diseases in this region. We identified 14 founder variants in Beauce that were not previously documented in Quebec, illustrating the pertinence of population genetics studies to document the increased risk of rare diseases for certain populations. The most frequent founder variant in Beauce, NM_000022.4:c.956_960del, associated with severe combined immunodeficiency, and variant NM_001698.3:c.656-2_656-1del, associated with 3-methylglutaconic aciduria type 1, were never described among French-Canadians despite high regional carrier rates. Additionally, two unreported founder variants were virtually absent from UrbanQc with carrier rates of 1/32 and 1/45 in Beauce. Interestingly, NM_001048174.2:c.1103G>A, one of the two most frequent variants associated with MUTYH-related cancer predispositions among individuals of Northern European descent^60^, was identified as founder in Beauce, but not UrbanQc, despite its spread through founder events that occurred during early European history^61^. It was, to our knowledge, never reported among individuals of French-Canadian descent.

While most founder variants are recessive, we identified three dominant founder variants, only one of which was previously reported. NM_007194.4:c.1100del is a frequently occurring variant in individuals of French-Canadian descent with a history of breast cancer^62^. The two other dominant founder variants were never reported in the French-Canadian population. NM_001039213.4:c.1186A>G, associated with rare genetic deafness, is globally rare and was found in 1/28 individuals in the Beauce cluster. It might have remained unnoticed in the region due to limited genetic testing for rare genetic deafness^63,64^. Similarly, NM_000552.5:c.7393G>A, associated with von Willebrand disease type 2 remained unnoticed despite its carrier rate of 1/45 in Beauce, possibly due to the challenges in this disease diagnosis^65^.

This study is limited by the sample size of the Beauce WGS cohort, which may not be large enough to accurately represent carrier rates across the entire general population of the region. As a result, our findings should be interpreted as a preliminary exploration rather than a definitive characterization. However, since individuals were not selected based on a rare disease diagnosis, and since the clustering was based on the genetic similarities between individuals, it could still enhance regional pathogenic variant discovery. While the results provide valuable insights into the rare variants enriched in the region, further studies with larger, more representative cohorts will be necessary to better estimate global carrier rates. Although, since the present-day Beauce population results from a regional founder effect, we might need a less important sample size to be representative of the population than for more admixed regions.

In conclusion, this study provides a genealogical and genetic characterization of Beauce, a regional population shaped by a limited number of founding families and geographic isolation. Those factors led to elevated kinship and inbreeding coefficients, a low ADR, and a unique genetic structure, as evidenced by a distinct cluster of individuals with Beauce ancestry in the UMAP visualisation of genotype data. Within this cluster, we identified 28 rare pathogenic founder variants, 14 of which were previously described in the province. This study highlights the demographic and genetic distinctiveness of Beauce and underscores the pertinence of fine-scale genetic analyses to uncover region-specific rare variants, even within a population that was previously thought to be homogenized by the initial founder effect. Our approach offers a powerful framework for the identification of rare variants and has potential to facilitate population health management. By focusing on smaller cohorts of individuals who share a common ancestry, this method enhances the discovery of rare variants as it increases their frequency. Moreover, applying this approach to existing large cohorts could uncover other populations with distinct genetic structures that may have been understudied, thereby enhancing the value of existing cohorts while remaining cost-effective^39,55,66^.

## 5 Data availability

The genealogical data is publicly available upon request via an independent data access committee by BALSAC (https://balsac.uqac.ca/acces-donnees/). The data from the Eastern Quebec schizophrenia and bipolar disorder kindred study is available upon request from MM. The data from the CARTaGENE cohort is publicly available via an independent data access committee by the CARTaGENE cohort (https://cartagene.qc.ca/en/researchers/access-request.html).

## 6 Author Contributions

MG, CM, SLG designed this study. MG analyzed the data and wrote this manuscript. AB and MM collected the samples. JR provided biostatistics expertise. M-CB provided technical assistance in the sample preparation. All authors contributed to the article and approved the submitted version.

## 8 Acknowledgments

The Eastern Quebec schizophrenia and bipolar disorder kindred study was funded by a Canada Research Chair (#950-200810) in psychiatric genetics of which MM is the Chair and by Canadian Institutes of Health Research grants (#MOP-74430, #MOP-114988, #PCG-155471, and #PJT-175122). Funding for SLG was provided by the Canada Research Chair (#CRC-2022-00444) in Genetics and Genealogy. MG received scholarships from the Fonds de recherche du Québec – Santé and the Canadian Institutes of Health Research.

