## Supplementary Material for "Rare variants and founder effect in an understudied Quebec population"

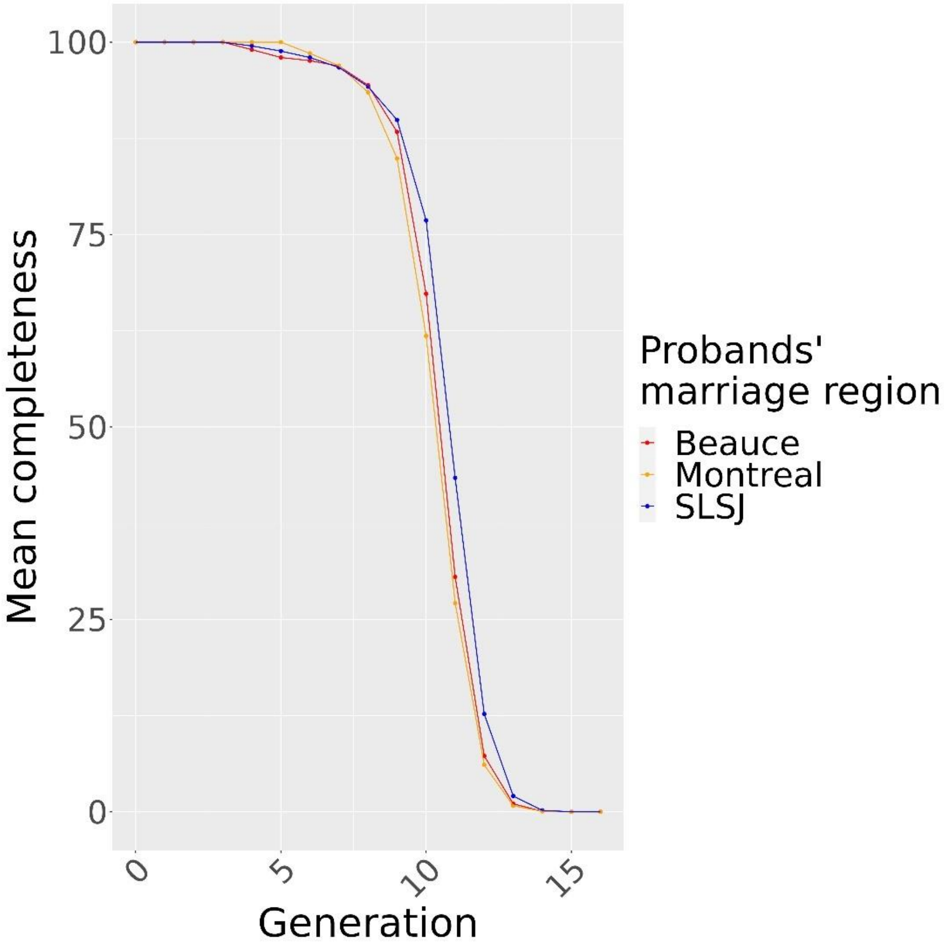


**Supplementary Figure S1.** Mean completeness per generation for each regional group

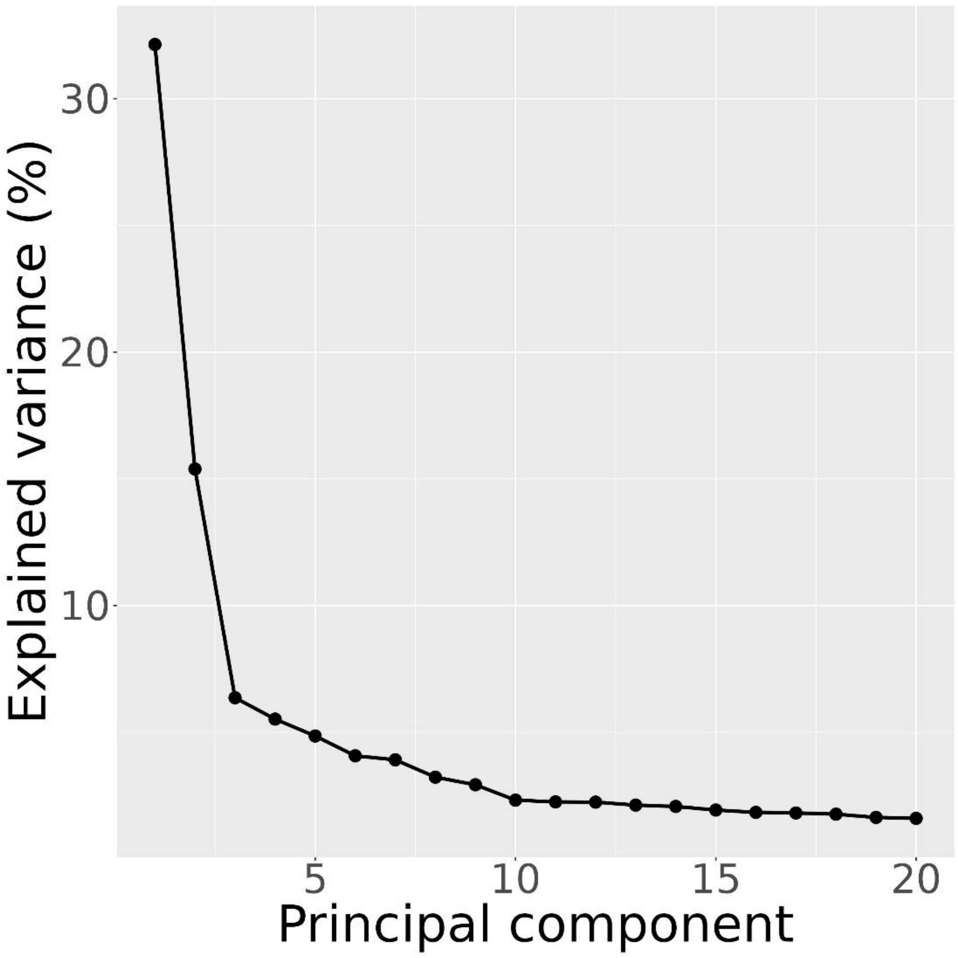


**Supplementary Figure S2**. Scree plot for the principal component analysis of the genotype data

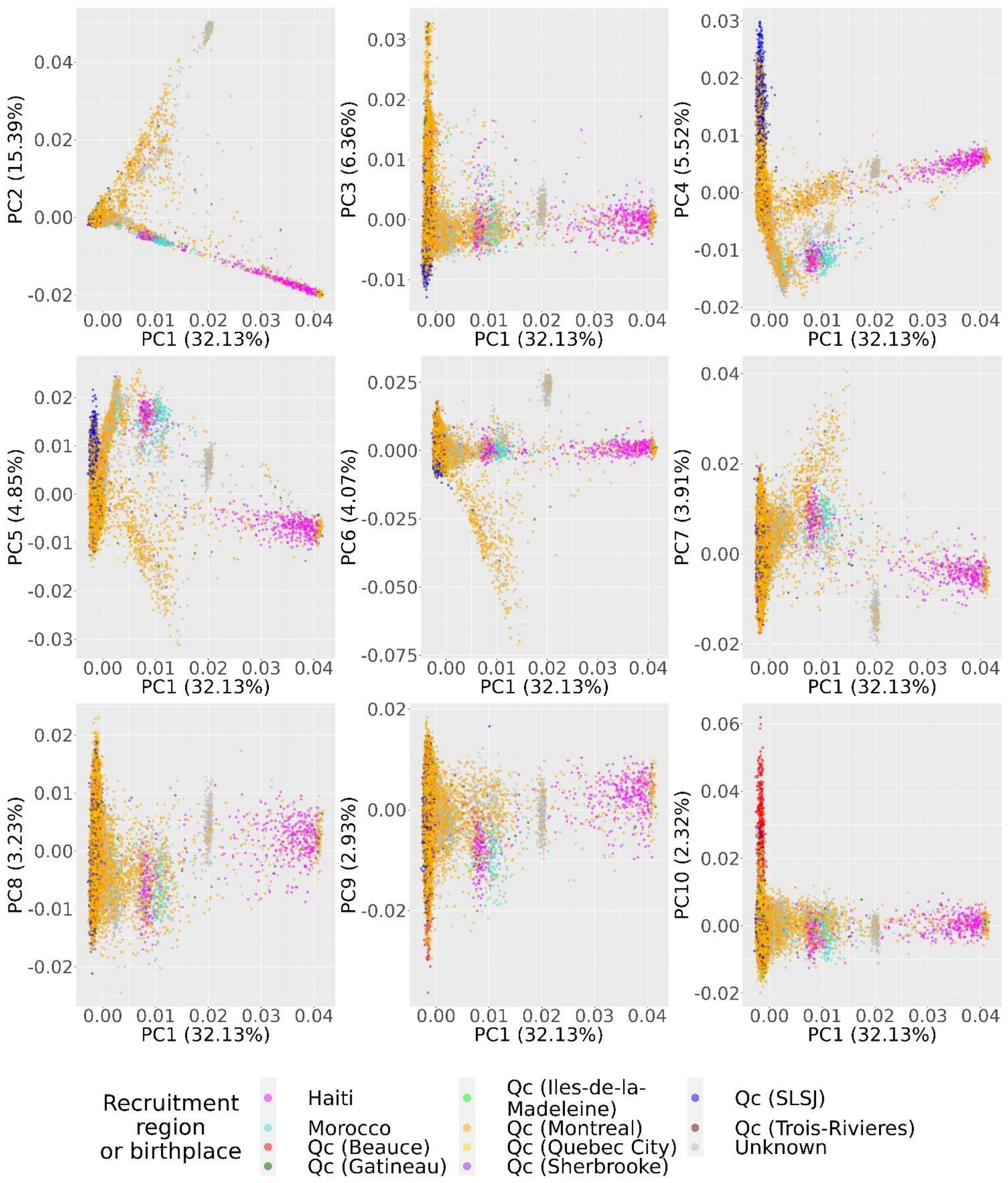


**Supplementary Figure S3.** Principal component analysis of the genotype data with individuals colored by recruitment region or birthplace if outside of Canada

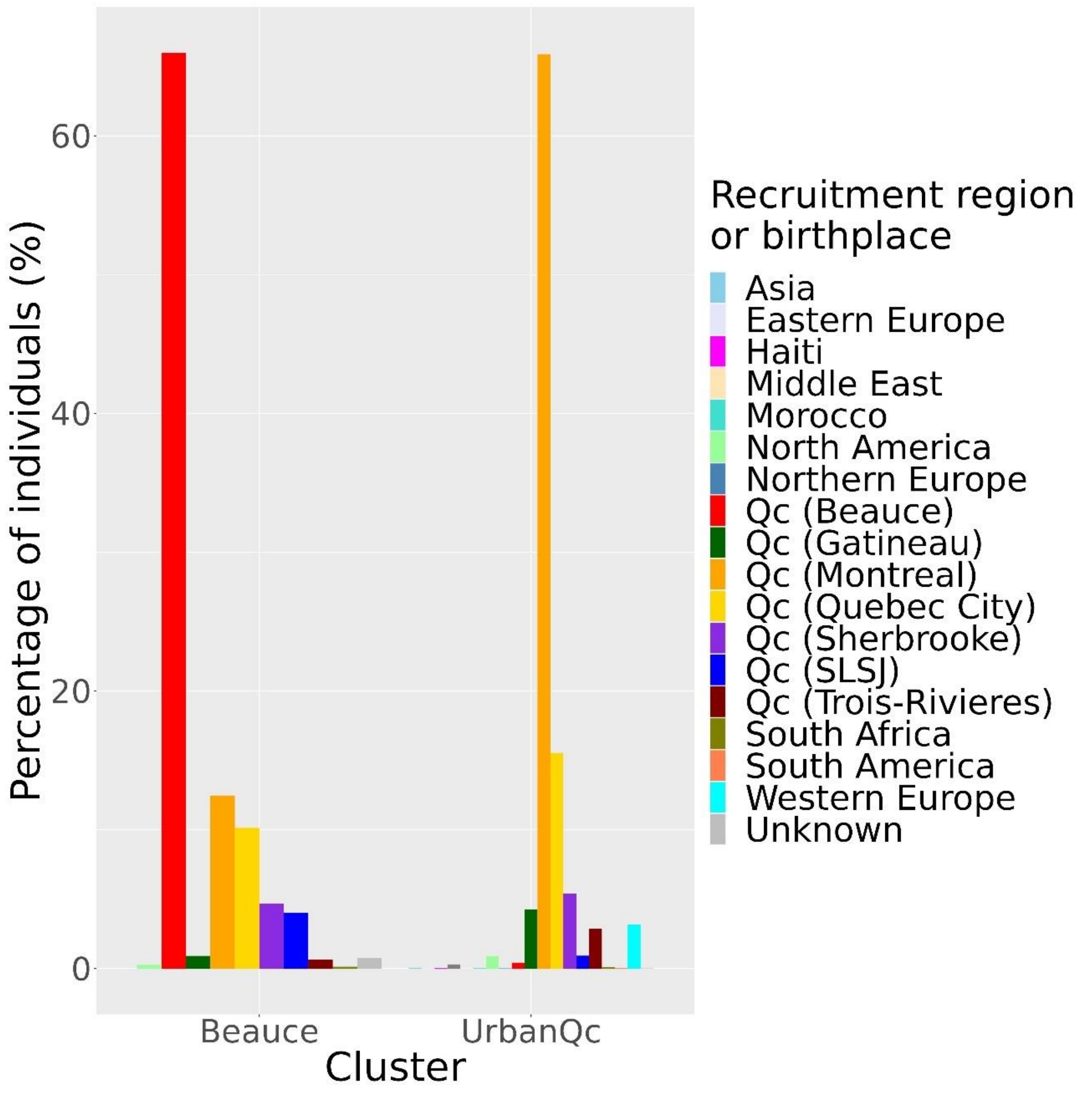


**Supplementary Figure S4.** Recruitment region or birthplace of individuals within the Beauce and UrbanQc clusters identified with DBSCAN on the UMAP performed on the 10 first principal components of the genotype data

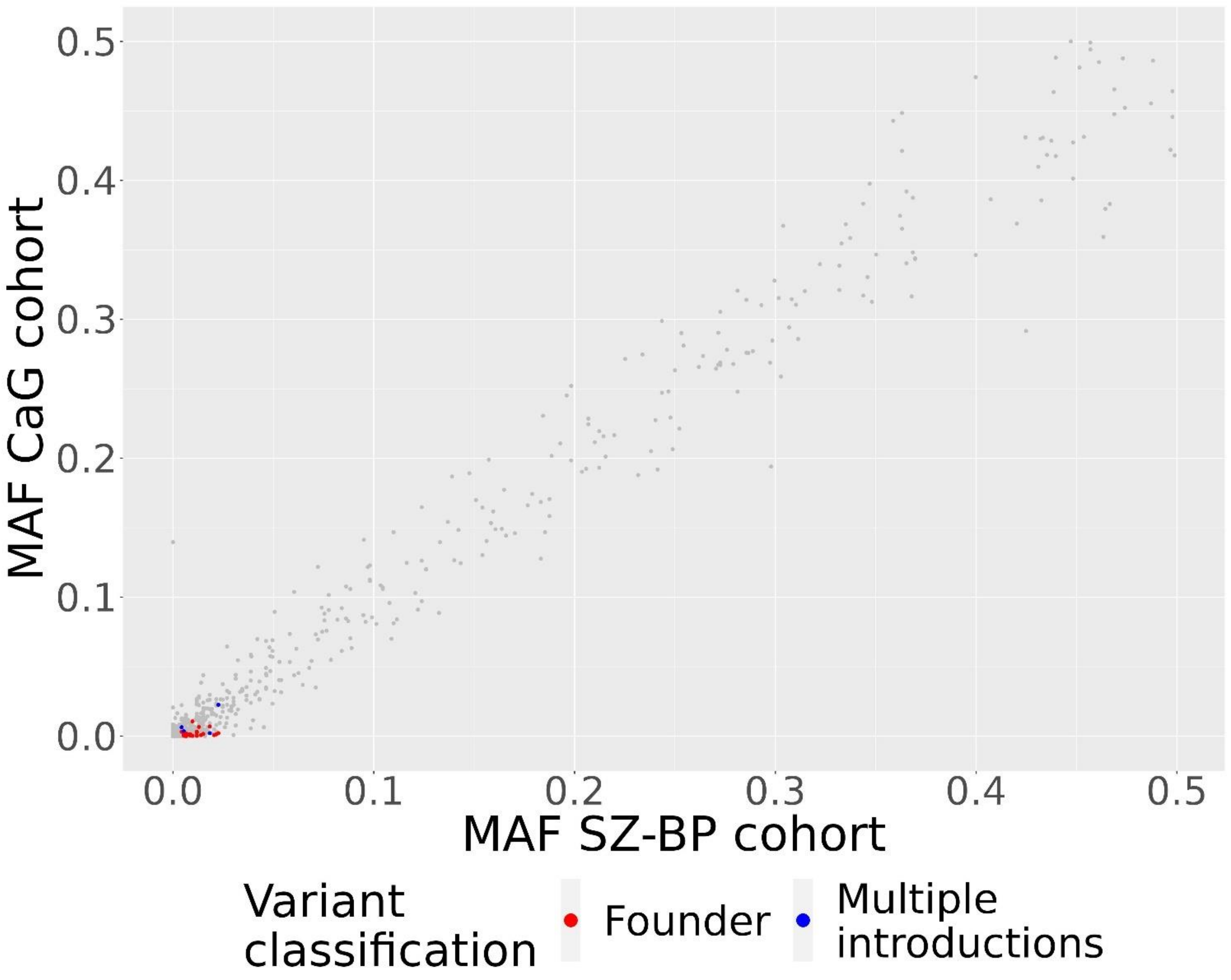


**Supplementary Figure S5.** Minor allele frequencies of the rare pathogenic variants in the SZ-BP cohort compared to the CaG cohort. Rare pathogenic variants with a relative frequency difference of at least 10% in Beauce compared to UrbanQc are highlighted

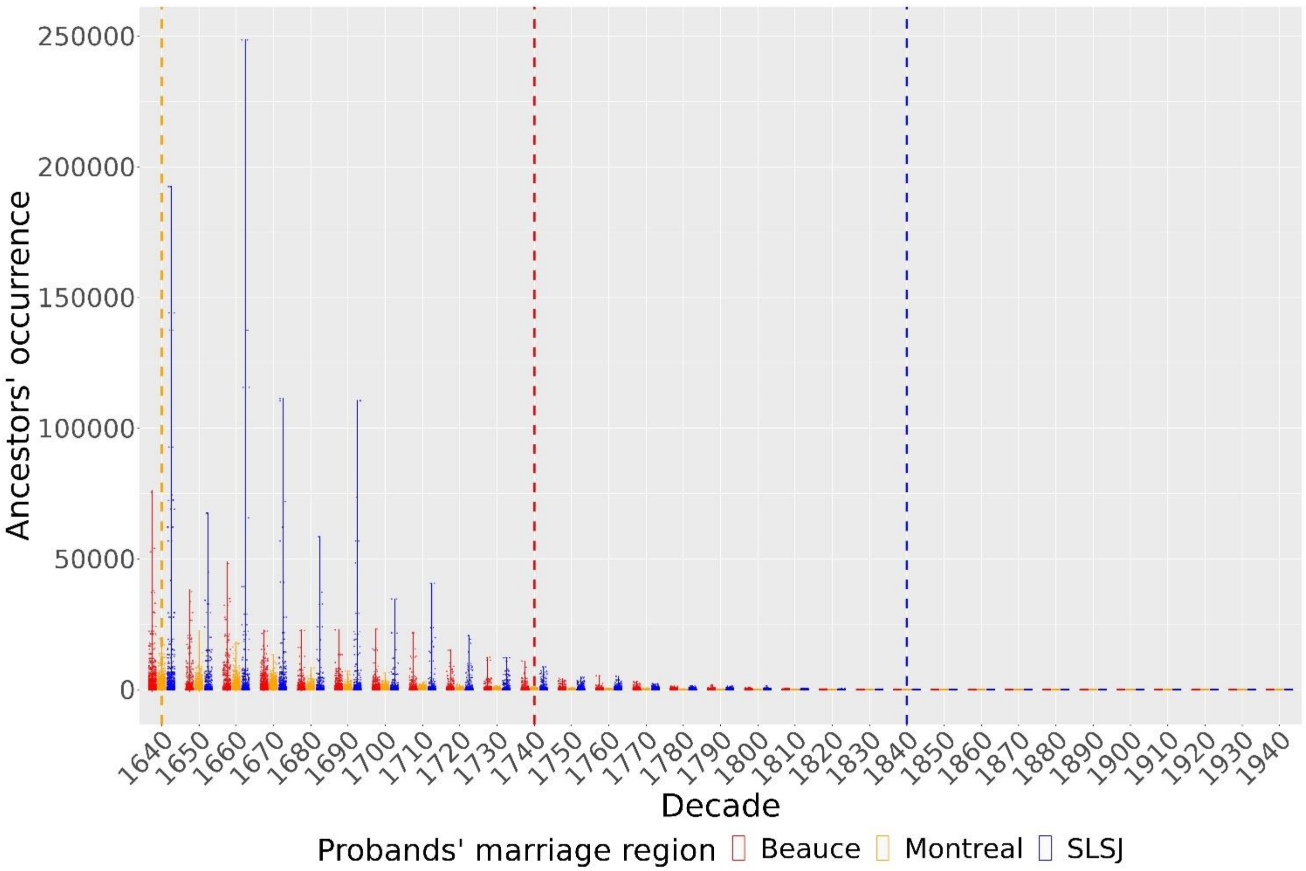


**Supplementary Figure S6.** Violin plot of the ancestors' occurrence of each regional group per decade. Vertical dotted lines represent the decade of the beginning of the settlement for each region

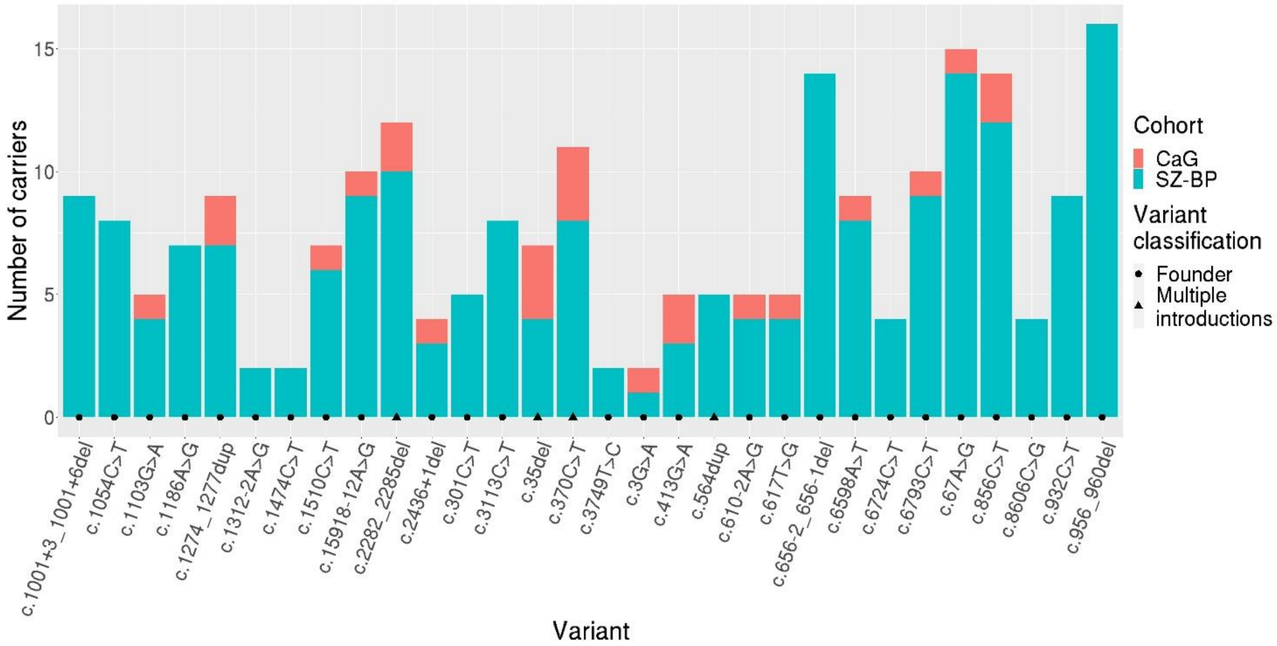


**Supplementary Figure S7.** Number of carriers in the Beauce cluster for each rare pathogenic variants with a relative frequency difference of at least 10% in Beauce compared to UrbanQc, categorized by cohort ($n_{CaG}$ = 26, $n_{SZ\text{-}BP}$ = 197)

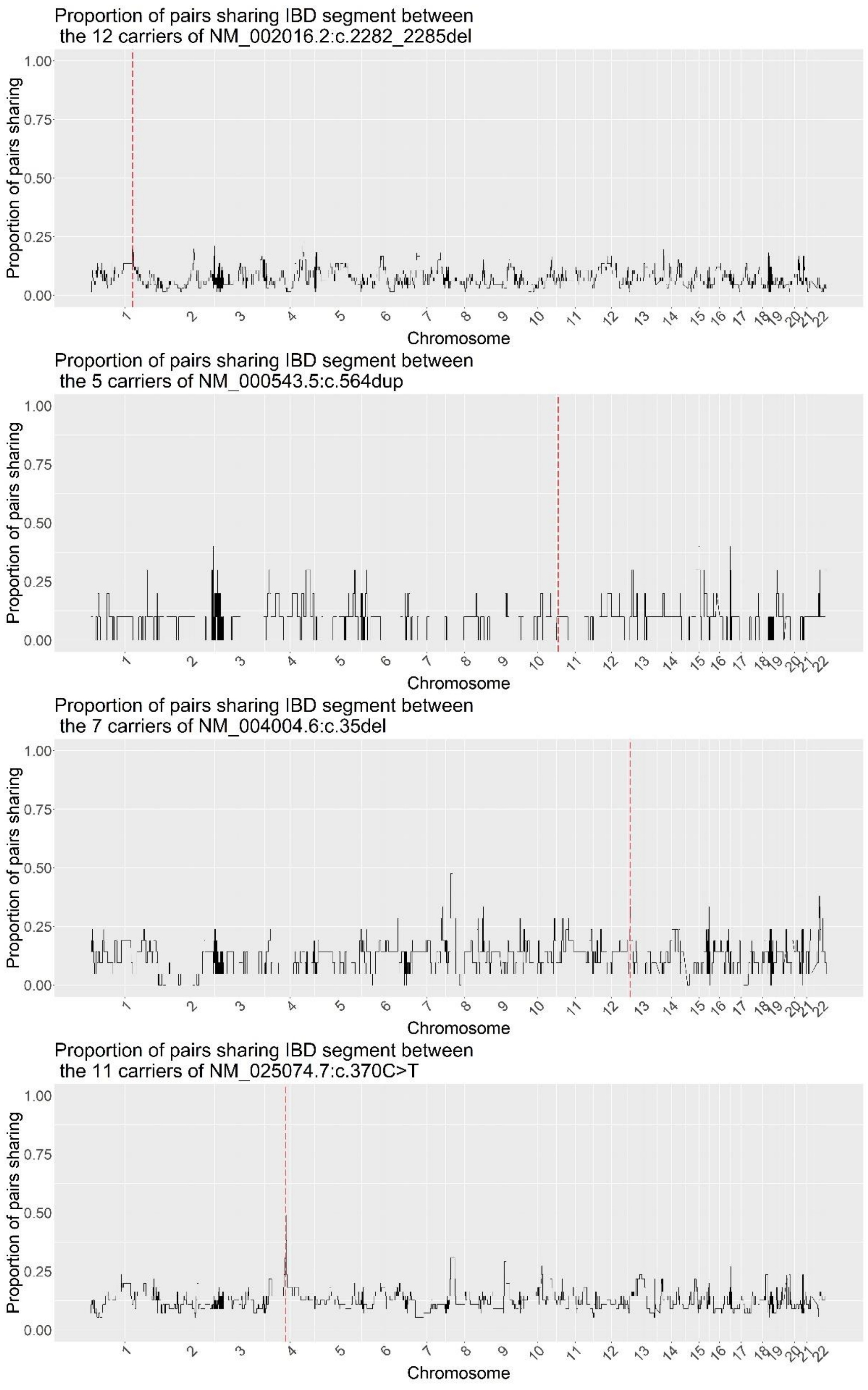


**Supplementary Figure S8.** Proportion of pairs of carriers sharing at least one haplotype IBD at each genomic position for the three variants with multiple introductions

**Supplementary Table S1.** Rare pathogenic variants with a relative frequency difference of at least 10% in Beauce compared to UrbanQc due to multiple introductions

| Gene | HGVS name | Disease (ClinVar ID) | CR Beauce | CR UrbanQc | MAF NFE |
|---|---|---|---|---|---|
| *FLG* | NM_002016.2:c.2282_2285del | Skin disorders and atopic conditions (16320) | 1/19 | 1/24 | 0.0233 |
| *FRAS1* | NM_025074.7:c.370C>T | Fraser syndrome type 1 (197861) | 1/20 | 1/109 | 0.0001 |
| *GJB2* | NM_004004.6:c.35del | Hearing loss and related disorders (17004) | 1/32 | 1/78 | 0.0083 |
| *SMPD1* | NM_000543.5:c.564dup | Niemann-Pick disease type 1 or 2 (632992) | 1/45 | 1/156 | 0.0039 |

CR: Carrier rate, MAF: Minor allele frequency, NFE: Non-Finnish Europeans
